# Mechanistic Insights into Skin Sympathetic Nerve Activity Dynamics in Healthy Subjects Through a Two-Layer Signal-Analytical and Closed-Loop Physiological Modeling Framework

**DOI:** 10.64898/2026.04.11.26350680

**Authors:** Runwei Lin, Frank R. Halfwerk, Dirk W. Donker, Jacomine Tertoolen, V.R. van der Pas, Gozewijn Dirk Laverman, Ying Wang

## Abstract

**Objective:** Skin sympathetic nerve activity (SKNA) has emerged as a promising non-invasive surrogate measure of sympathetic drive, but its relevant physiological characteristics remain ill-defined. This observational study aims to investigate its regulatory patterns during rest and Valsalva maneuver (VM) in healthy participants.

**Method:** Using a two-layer strategy integrating signal analysis and physiological modelling, we analyzed data recorded from 41 subjects performing repeated VMs. The observational layer includes time-domain feature comparisons using linear mixed-effect models, and time-varying spectral coherence analysis. The mechanistic layer proposes a mathematical model to investigate whether baroreflex and respiratory modulation are sufficient to reproduce the observed HR and average SKNA (aSKNA) dynamics.

**Main Results:** Mean integrated SKNA (iSKNA) showed more significant change than HRV for VM induced effects. We also found mean iSKNA increase during VM varies with BMI and sex. The coherence analysis indicated that iSKNA strongly synchronized with EDR under resting conditions. The proposed model successfully reproduced main characteristics of aSKNA dynamics, yielding a high median Pearson correlation coefficient of 0.80 ([Q_1_, Q_3_] = [0.60, 0.91]). In contrast, HR dynamics were only partially captured, with a median PCC of 0.37 ([Q_1_, Q_3_] = [0.16, 0.55]). These results likely suggest SKNA provides a more direct representation of sympathetic burst dynamics during VM in healthy subjects.

**Significance:** This study provides convergent evidence that SKNA reflects known autonomic regulatory influences in healthy subjects. These findings strengthen the physiological interpretability of SKNA while clarifying its appropriate use as a practical biomarker of sympathetic function.

## I. Introduction

Skin sympathetic nerve activity (SKNA) has recently been proposed as a non-invasive biomarker for sympathetic nervous activity. It has shown to be extractable from the high-frequency components of the electrocardiography (ECG) [1]–[3] and may be more sensitive than traditional markers such as electrodermal activity [4] and heart rate variability (HRV) [5] in detecting sympathetic activation during acute autonomic challenges, such as Valsalva maneuver (VM). Since its first introduction [2], SKNA has been applied in multiple autonomic and cardiovascular applications, for instance, obstructive sleep apnea [6], [7], cardiac arrhythmias [8], [9], and the characterization of pain and emotional stimuli [4].

Despite these promising and clinically relevant applications, the regulatory mechanism of SKNA remains incompletely understood. Observed SKNA fluctuations may arise from multiple physiological influences rather than a single regulatory mechanism. For example, increasing SKNA during autonomic challenge are consistent with sympathetic activation [2], [4], while spectral analysis during sleep has reported integrated SKNA power components aligned with the respiratory frequency, suggesting respiratory-related modulation [7]. These observations suggest that SKNA may reflect the superposition of multiple physiological mechanisms whose relative contributions may vary across states. A similar complexity has been recognized in traditional invasive sympathetic nerve recordings obtained using microneurography, where skin (SSNA) and muscle (MSNA) sympathetic activities can reflect different regulatory processes depending on the experimental context [10]. Nevertheless, existing studies of SKNA focused primarily on demonstrating potential clinical applications of SKNA, while the physiological drivers underlying its observed dynamics remain insufficently validated. This limits its interpretation as a quantitative biomarker of sympathetic activity.

This observational study aimed to assess the sensitivity of SKNA to sympathetic activation, and to validate its regulatory patterns during rest and the Valsalva maneuver (VM) in healthy participants. We hope to provide evidence to delineate the conditions under which SKNA can be meaningfully interpreted as a state-dependent marker of sympathetic activity. To address this gap, we proposed a two-layer validation framework that integrates observational signal analysis including linear mixed-effects model (LMM) and time-spectral analysis, with mathematical modelling. A preliminary version of this model was presented before [11]. The present study substantially redesigns the model and extends it by providing combined signal-level and model-based validation of SKNA dynamics during VM.

## II. Method

This study includes a two-layer complementary validation. Within the obeservational layer of our proposed analysis framework, we first characterized the basic properties of iSKNA by comparing its changes with time domain HRV during VM and resting periods. Second, we analyzed the timevarying power of iSKNA and HRV in low frequency (LF) and high frequency (HF) bands. Considering the respiratory impact, we further analyzed the time-varying coherence between iSKNA and ECG-derived respiration (EDR). Within the mechanistic layer, we developed a simplified closed-loop mathematical model incorporating baroreflex control and respiratory modulation to explain the observed iSKNA dynamics and to jointly simulate HR and SKNA responses during VM.

### A. Ethics approval and data description

Initial data collection in healthy volunteers was approved by the Natural Science and Engineering Sciences Ethics Committee of the University of Twente (2022.153). Informed consent was signed by all volunteers. The reuse of the data for this study was approved by the Ethics Committee of the Computer and Information Sciences, University of Twente (nr. 230713). Data from 41 healthy subjects were collected at the lab located at Thorax Centrum Twente, Medisch Spectrum Twente, the Netherlands. Each participant underwent two measurements. For each measurement, participants were asked to perform two VM sessions, each lasting 15 seconds in a lying position. The dataset used in this study was previously described elsewhere [11] and will be published at Zenodo (doi: 10.5281/zenodo.19116774).

Simultaneous recordings of ECG and SKNA (neuECG) were recorded using a Biomonitor (Mega Electronics Ltd., Kuopio, Finland) at a sampling frequency of 9600 Hz. ECG electrodes were positioned according to the Einthoven triangle configuration, with lead I used for the first measurement and lead III for the second, chosen for their superior signal-to-noise ratio. For SKNA extraction, the neuECG signals were bandpass filtered between 500 and 1000 Hz using a FIR filter with 100 dB attenuation at the cutoff frequencies, as previous study reported that this frequency range is optimal for SKNA analysis [1]. The high-frequency signals were then rectified and integrated using a moving average method by a 100-ms window to obtain integrated SKNA (iSKNA). For HR estimation, signals were bandpass filtered between 1–40 Hz with a 400th-order FIR filter to remove baseline wander and high frequency noise. R peaks were then detected using the Pan–Tompkins algorithm [12]. Among the 82 measurements (41 measurements × two measurements), one was excluded due to technical failure and two due to invisible or unreliable R peaks during the VM, leaving 79 measurements and 158 VM sessions from 41 subjects for analysis.

Because iSKNA is highly spiky and stochastic, which complicates dynamical modelling, we chose to model average SKNA (aSKNA) instead of iSKNA. The aSKNA signal was derived by applying a 1-s moving average filter to the iSKNA, yielding a smoother waveform. The HR, iSKNA and aSKNA were resampled to 10Hz using the piecewise cubic Hermite interpolating polynomial for alignment. The aSKNA preserves the slow dynamics and VM-induced changes observed in iSKNA, with highly similar temporal profiles. As each measurement containing two VM sessions, each measurement was divided into two segments, with the first 90 s assigned to the first VM session and the remaining 85 s to the second VM session. Fig. 1 shows the 50% trimmed mean (mean value of data between Q_1_ and Q_3_) and interquartile range (IQR) of the derived HR, iSKNA and aSKNA from 79 measurements.

**Fig. 1.**
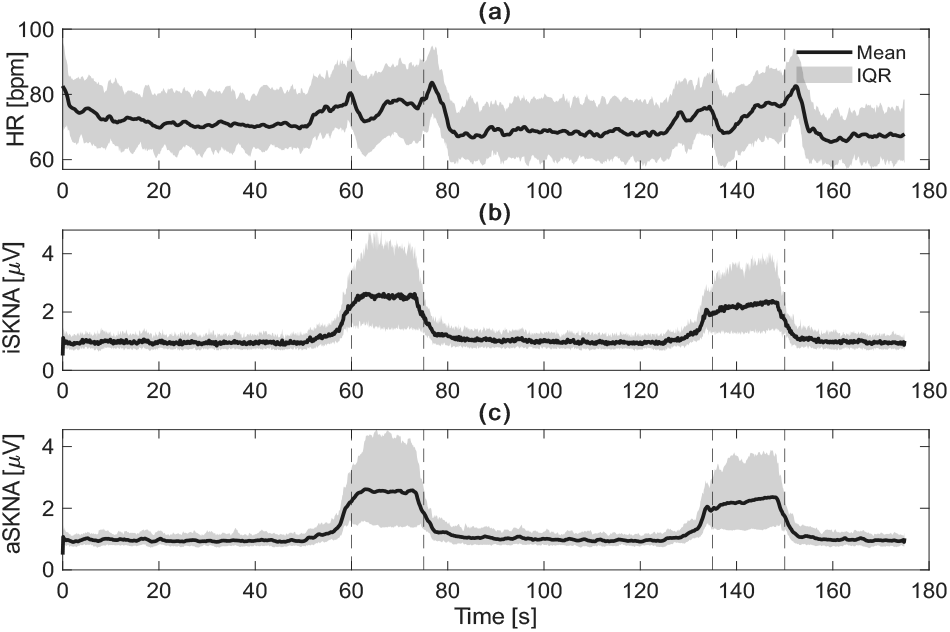
50% Trimmed Mean and interquartile range (IQR) of derived signals is displayed (a) Heart rate (HR); (b) integrated SKNA (iSKNA); (c) average SKNA (aSKNA). Onset and end time of Valsalva maneuvers were labelled by vertical lines. Each measurement was divided itnto two sessions for model validation. SKNA = Skin sympathetic nerve activity.

Respiration induces rhythmic oscillations in intrathoracic pressure (ITP), which mechanically modulates heart rate [13]– [15]. It is therefore essential to account for respiratory influences in any analysis of autonomic cardiovascular regulation. We adopted a state-of-the-art method based on the QRS complex area [16], [17] to estimate ECG-derived respiration (EDR). Briefly, the ECG was firstly bandpass filtered between [5Hz, 35 Hz] by a fourth order Butterworth filter to suppress components other than QRS complex. For each beat, the onset and offset of the QRS complex were identified using zero-crossings of the ECG derivative, and the QRS area was calculated accordingly.

### B. Linear mixed model analysis on time domain features

We employed LMM to assess difference in mean iSKNA and HRV between VM and resting periods (State) to reproduce previously reported autonomic responses [2], [4]. LMMs were chosen given the hierarchical data structure arising from repeated measurements within subject and inter-measurement variability, and the potential confounding factors, including age, sex and BMI, given their known impact on ANS [18], [19]. We first checked the effects of different factors separately using nested models. Detailed results of these nested models are summarized in supplementary material I. BMI and age were centered by subtracting their mean values before analysis.

The final model is defined as:

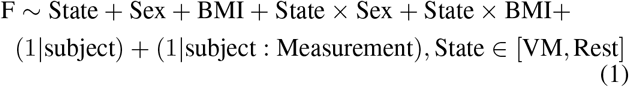

where F refers to the physiological metric of interest: either mean iSKNA or the root mean square of successive differences (RMSSD) of RR intervals, which is a conventional marker of short-term vagal modulation [20]. For each measurement, metrics were calculated from two VM periods and two resting period separately. Within the LMM, we used type III F tests with Satterthwaite’s method to investigate the effect of VM (noted by the variable State in the LMM model) on SKNA and HRV. A p-value of 0.05 was deemed significant. The analysis was performed with the lmer function from lme4 package in R software [21].

### C. Time-varying spectral analysis

Time-varying spectral analysis is widely used in analyzing autonomic cardiovascular regulation under unstable conditions [22], [23], as different frequency bands in cardiovascular oscillations correspond to specific physiological processes: respiratory related modulation stays mostly in HF (0.15Hz-0.4Hz), and LF (0.04Hz-0.15Hz) reflect a mixture of baroreflex-mediated autonomic modulation (sympathetic and parasympathetic) together with slower hemodynamic and vascular regulatory mechanisms [20]. Therefore, this study first performed a time-frequency analysis to examine the time-varying spectral properties of iSKNA, RR interval series, and EDR. We employed the multitaper spectrogram (MTSP), a method well-validated for analyzing non-stationary cardiovascular oscillations [22]. The MTSP in this study was implemented based on the package introduced in [24]. Computation of the spectrogram uses a window length of 25 seconds and stride of one second. While the 25-s window length is longer than the VM duration, it is chosen for the a minimum differentiable frequency of 0.04Hz, which is the lower bound of LF components. The spectrogram was averaged over three discrete prolate spheroidal sequence tapers, which is a commonly used orthogonal window. Consequently, the band power was integrated in LF and HF range, respectively. The time-varying coherence between two time series *x* and *y* (iSKNA-EDR or RR–EDR in our analysis) was calculated as:

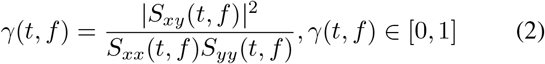

where *S*_*xy*_(*t, f* ) is the cross-spectrum between *x* and *y* at frequency f and time t. *S*_*xx*_(*t, f* ) and *S*_*yy*_(*t, f* ) are the self-spectrum of two series *x* and *y*, respectively. We studied the time-varying coherence between iSKNA and EDR, as well as between iSKNA and RR series. The band coherence was calculated by averaging the *γ*(*t, f* ) over the LF and HF frequency range, respectively.

### D. Mathematical model of aSKNA and HR dynamics during VM

#### 1) Model assumptions and descriptions

We built a mathematical model that explicitly incorporates both baroreflex and respiration modulation, to validate the possible aSKNA and HR regulation mechanisms. This model was developed based on the following assumptions:

1. HR and aSKNA dynamics during VM are assumed to be predominantly governed by arterial baroreflex and respiratory modulation, while other metabolic and mechanical influences have minimum influence. This assumption is motivated by the observations from signal analysis: the data exhibit clear sympathetic arousal during VM and respiration-related modulation under resting period.
2. The cardiovascular system is represented by a simplified three-compartment structure consisting of the venous compartment, the heart, and the arterial compartment. This was down both to maintain model tractability and given the limited observation data.
3. We simplified the medulla autonomic integration by representing it as a direct mapping from baroreceptor firing rate to sympathetic and parasympathetic efferent activity.
4. During the strain phase of VM, subjects were assumed to generate an intrathoracic pressure (ITP) plateau of approximately 35 mmHg, and normal respiratory influence was considered negligible. This assumption was done because direct ITP measurement was not available in our recordings. It is based on the standardized VM protocol reporting typical ITP levels of 30–40 mmHg [25].

Fig. 2 illustrates the architecture of the proposed model, which comprises an autonomic compartment and a cardiovascular compartment. The autonomic compartment includes the baroreceptor afferent and efferent pathways, while the cardiovascular compartment is simplified as the three components mentioned above.

**Fig. 2.**
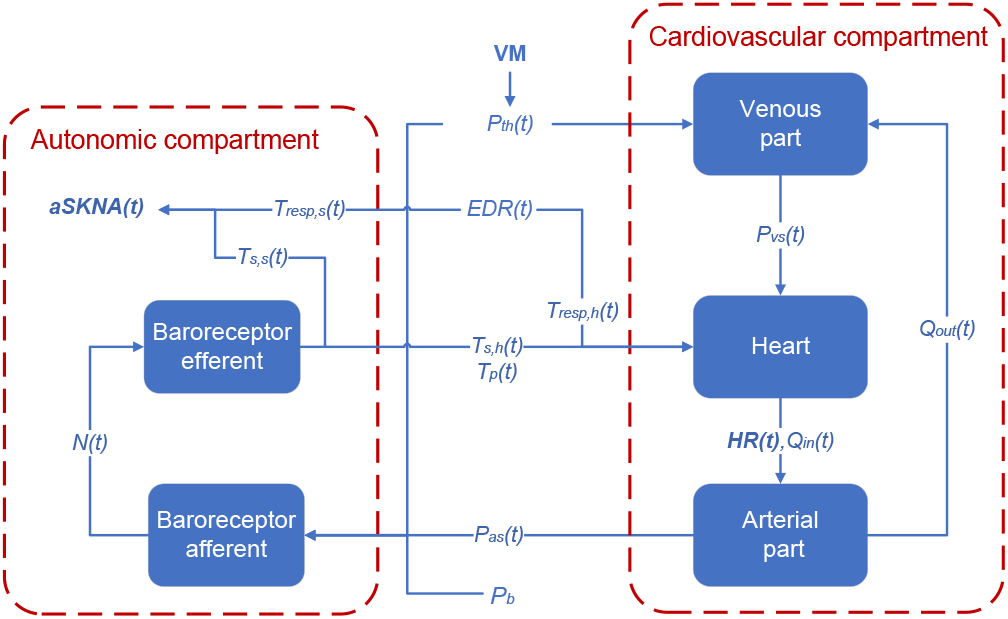
Scheme of model architecture. The closed-loop model incorporated an autonomic and a cardiovascular compartment. Q_out_**(*t*)** = blood flow out of arterial part into venous part; P_***vs***_**(*t*)** = venous pressure; HR(t) = heart rate; Q_in_**(*t*)**= cardiac output; P_***th***_**(*t*)** = intrathoracic pressure; EDR(t) = ECG-derived respiration; T_resp,h_**(*t*)** = respiration modulated heart rate related parasympathetic tone; T_***s***,***h***_**(*t*)** = baroreflex modulated heart rate related sympathetic tone; T_***p***_**(*t*)** = baroreflex modulated heart rate related parasympathetic tone; P_as_**(*t*)** = arterial blood pressure; P_***b***_ = baseline blood pressure; T_resp,s_**(*t*)** = respiration modulated aSKNA related sympathetic tone; T_***s***,***s***_**(*t*)** = baroreflex modulated aSKNA related sympathetic tone; ***N* (*t*)** = baroreceptor firing rate. VM = Valsalva maneuver.

As mechanistic modelling of SKNA dynamics requires the presence of clear sympathetic activation events, only VM sessions exhibiting pronounced SKNA response (bursts) were included for model validation. Accordingly, a burst analysis was performed for each VM session to check whether sufficient bursts present. For the burst threshold estimation, each iSKNA recording was modelled as a mixture of two Gaussian distributions using the Expectation–Maximization algorithm. The burst threshold was defined as the mean of the lower Gaussian plus three times its standard deviation, in line with the method proposed in [6]. A session was included if more than 30% (empirical threshold) of the iSKNA amplitudes during VM exceeded this threshold, i.e., clear SKNA burst observed. In total, 125 out of 158 VMs (79.1%) met the criteria and were used for model validation.

#### 2) Intrathoracic pressure input during VM

VM introduces a sustained ITP elevation, which alters venous return and triggers a cascade of cardiovascular responses. In this study, the ITP *P*_*th*_(*t*) [mmHg] was modeled as a sustained 35 mmHg pressure during the VM. To avoid discontinuities at the onset and offset of VM and prevent numerical stiffness, we used a smoothed waveform by combining two hyperbolic tangent (tanh) functions:

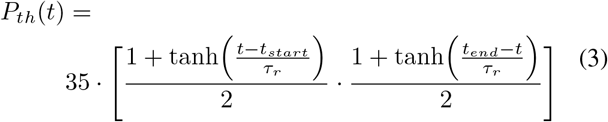

where *τ*_*r*_ [s] is a time constant controlling the transition speed at VM onset *t*_*start*_ [s] and offset *t*_*end*_ [s], and was set to 0.6s empirically in this study.

During VM, the elevated ITP impedes peripheral venous return. We assumed this effect can be modeled by the following first-order differential equation (ODE) for venous pressure *P*_*vs*_(*t*):

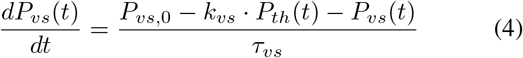

where *P*_*vs*,0_ denotes the baseline venous pressure, *k*_*vs*_ is a gain factor describing ITP’s impact on venous pressure reduction, and *τ*_*vs*_ is a time constant that decides the speed of venous pressure adaptation.

#### 3) Cardiovascular model

A simplified closed-loop model was employed to describe cardiovascular responses to the altered venous return. The change of arterial systolic BP (SBP) *P*_*as*_(*t*) [mmHg] was described by a first-order Windkessel model, following [26]:

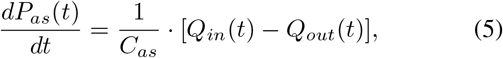

where *C*_*as*_ [L/mmHg] was arterial compliance, *Q*_*in*_(*t*) was cardiac output, and *Q*_*out*_(*t*) [L/s] denotes arterial outflow. Cardiac output was defined as the product of stroke volume and HR:

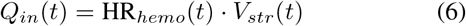

where the stroke volume *V*_*str*_(*t*) was modeled as a ratio of venous to arterial pressure scaled by the contractility coefficient *k*_*str*_ [mmHg/beat] according to [26]:

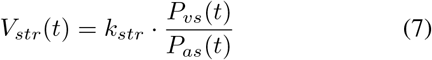

The calculation of *k*_*str*_ is summarized in supplementary material II.

Arterial outflow *Q*_*out*_(*t*) was calculated using Ohm’s law:

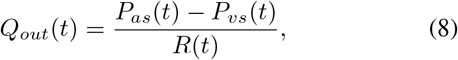

with total peripheral resistance *R*(*t*) was expressed as a linear function of sympathetic activity *T*_*s,h*_(*t*) [26]:

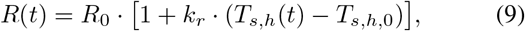

where *R*_0_ [mmHg · s/L] denotes baseline resistance, *k*_*r*_ denotes the sympathetic gain, and *T*_*s,h*,0_ represents the baseline sympathetic tone.

#### 4) Afferent baroreflex model

During VM, sustained ITP *P*_*th*_(*t*) changes the venous blood flow and perturbs SBP *P*_*as*_(*t*), consequently alters baroreceptor firing rate *N* (*t*) [Hz] and modulates autonomic tone through the baroreflex pathways. In this study, the change in baroreceptor firing rate was modeled by the ODE used in [26]–[28]:

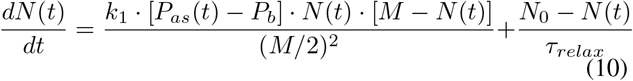

where *P*_*b*_ was the baseline SBP, *M* was the maximum firing rate, *N*_0_ represented the baseline firing rate. *τ*_*relax*_ and *k*_1_ [Hz/mmHg· s] were parameters that determine the response amplitude. This formulation produces the characteristic pressure–response of baroreceptors: N(t) increases when *P*_*as*_(*t*) rises, and decreases when *P*_*as*_(*t*) falls.

#### 5) Efferent baroreflex model

The efferent model describes the baroreflex response to the autonomic tone, which further affects the aSKNA and HR dynamics. The sympathetic and parasympathetic were thus modelled based on the baroreceptor firing rate separately. These efferent autonomic tones were modeled in two steps [29]:

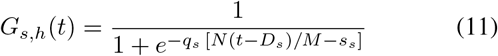

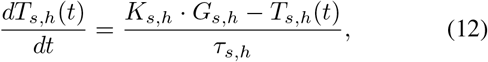

where *q*_*s*_ determined the steepness and *s*_*s*_ was the half saturation point, *K*_*s,h*_ was the scaling constant, and *τ*_*s,s*_ [s] was adaptation time constant. A empirical sympathetic delay was commonly assumed in previous models given the known delayed effect in cardiac sympathetic innervation [27], [29], [30]. In this study, we utilized a fixed sympathetic delay *D*_*s*_=3s, which is in line with [29], [30].

The parasympathetic response is mediated by the nucleus ambiguus and conveyed via vagal efferents to the sinoatrial node. Therefore, vagal conduction is much faster than the sympathetic innervation, and the parasympathetic delay can be considered neglectable [29]. The parasympathetic tone *T*_*p*_(*t*) was modeled in a similar way to *T*_*s,h*_(*t*) but with no delay:

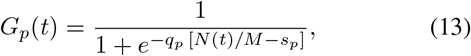

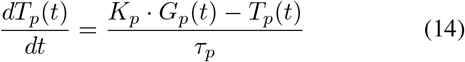

where *q*_*p*_ is the steepness factor and *s*_*p*_ was the half saturation point, *K*_*p*_, was the scaling constant, and *τ*_*s,p*_ [s] was the adaptation time constant.

The skin sympathetic pathway reflected by SKNA was also modeled without delay, as it origins from sympathetic nerves from post stallate ganglion and has fast kinetics. Here, we modelled the SKNA related sympathetic tone *T*_*s,s*_(*t*) as

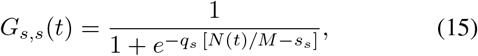

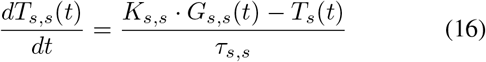

where *q*_*s*_, *s*_*s*_, and *K*_*s,s*_ were shared parameters with *T*_*s,s*_(*t*), *τ*_*s,s*_ [s] is the time constant which is smaller than *τ*_*s,h*_ [s] considering aSKNA have a quicker response. The calculation of the half-saturation values were summarized in supplentary material II.

#### 6) Respiratory modulation of HR and Askna

Respiratory influences were modelled as an effective autonomic modulation based on EDR under resting conditions, reflecting established physiological coupling between respiratory rhythm and autonomic outflow [31]. We assumed the respiratory modulation only existed during resting and was depressed during the VM due to voluntary breath holding, leaving baroreflex-mediated mechanisms as the dominant drivers of HR and SKNA dynamics. Accordingly, the respiratory modulation of HR and aSKNA were developed based on EDR in this study. The extracted EDR was normalized to zero mean and unit variance firstly. As RSA does not predominate during the VM, an attenuating window *W* (*t*) was applied to the EDR during the VM period. The RSA-related autonomic tone *T*_*resp,k*_(*t*) were then modeled as:

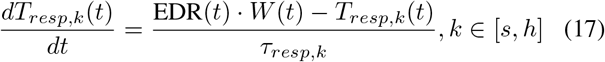

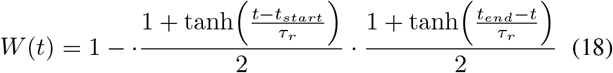

where EDR(*t*) denotes the normalized EDR and *τ*_*resp,k*_ [s] is the time constant representing the adaptation speed of *T*_*resp,k*_(*t*). This first-order ODE smooths the respiratory input. The respiratory modulation was modeled separately for RR (*k* = *h*) and aSKNA (*k* = *s*).

#### 7) HR and aSKNA estimation

In this study, HR was assumed to be modulated by both sympathetic and parasympathetic activity, with the latter including contributions from the both baroreflex and RSA. Accordingly, HR was modeled using the linear formulation proposed in [29]:

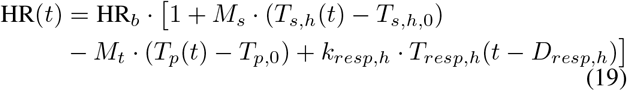

where HR_*b*_ [bpm] is the baseline HR and is defined as the median HR during resting period. *T*_*s,h*,0_ and *T*_*p*,0_ denote baseline sympathetic and parasympathetic tones, and were calculated with *N* (*t*)=*N*_0_ and Eq. 11-14. *D*_*rsa*_ represents the vagal delay associated with RSA.

SKNA arises from post-ganglionic sympathetic fibers from the stellate ganglion to the skin. Therefore, aSKNA was driven solely by the sympathetic tone without an additional delay using a linear equation:

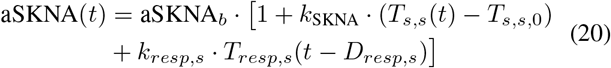

where aSKNA_*b*_ [*µ*V] is the baseline SKNA and is defined as the median aSKNA value from the baseline period. *T*_*s,s*_(*t*) the sympathetic activity relevant for skin, and *T*_*s,s*,0_ is its baseline value and equals to *T*_*s,h*,0_. *k*_SKNA_ is a proportionality coefficient describing the sensitivity of SKNA to changes in sympathetic tone. *k*_*resp,s*_ decides the respiratory influence on aSKNA, and *D*_*resp,s*_ represents the delay associated with respiratory modulation.

#### 8) Parameter identification and performance evaluation

As we assumed resting period is primarily modulated by respiration and VM period is mainly influenced by baroreflex, the model parameter identification follows a two-step approach: identification of respiratory modulation related parameters during resting period; and identification of model parameters during VM. We performed a sensitivity analysis to select a subset of parameters to be optimized considering the VM model contains a large number of parameters. Detailed description of the sensitivity analysis is summarized in supplementary material III.

Since respiration predominantly drives HR fluctuations during the resting period, the six relevant parameters *τ*_*resp,h*_, *k*_*resp,h*_, and *D*_*resp,k*_ were optimized only within this period. The parameters were optimized using the nonlinear leastsquares solver (function *lsqnonlin*.*m*) with the trust-region reflective method in MATLAB (MATLAB 2025a, The Mathworks, Natick, MA, USA), which is a widely used nonlinear optimization method [32]. The loss function was defined as the mean squared error (MSE) between the measured HR and aSKNA, and the model prediction for:

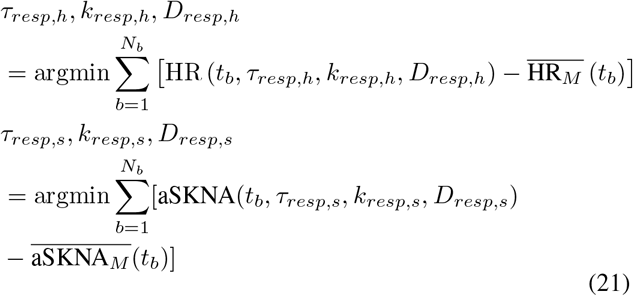

where *t*_*b*_ and *N*_*b*_ refers to the time and number of sampling points for the resting period where respiratory modulation dominates. 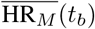 and 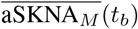 were measured HR and aSKNA at time *t*_*b*_.

We then performed a sensitivity analysis to select a subset of model parameters to be optimized. The selected parameter subset is: *θ*_*sub*_ = {*M, k*_*vs*_, *s*_*p*_, *R*_0_} (for details: supplementary material III). The selected parameters were then identified by minimizing the following loss function:

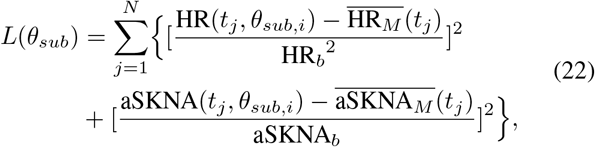

where 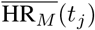 and 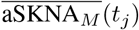 were measured HR and aSKNA at time *t*_*j*_, *θ*_*sub,i*_ denotes the ith parameter in the selected parameter subset *θ*_*sub*_. The residuals were normalized to remove scale differences between aSKNA and HR. The minimization was performed using the trust-region reflective algorithm. Other parameters were fixed at their nominal values, which are selected based on literature and empricially. Table I summarizes the fixed parameter values.

**TABLE I.** Summary of fixed parameters with their definitions, nominal values, and units.

| Parameters | Definition | Nominal value | Unit |
| --- | --- | --- | --- |
| $N_0$ | Baseline baroreceptor firing rate | 35 | Hz |
| $D_s$ | Sympathetic delay of HR control | 3 [29] | s |
| $s_{s,h}$ | Half-saturation value for sympathetic HR control | 0.03 | - |
| $s_{s,s}$ | Half-saturation value for sympathetic aSKNA control | 0.03 | - |
| $q_{s,h}$ | Steepness of sympathetic HR control | 10 [29] | - |
| $q_t$ | Steepness of parasympathetic HR control | 10 [29] | - |
| $q_{s,s}$ | Steepness of sympathetic aSKNA control | 10 | - |
| $K_{s,h}$ | Gain of baroreflex sympathetic control of HR | 5 | - |
| $K_p$ | Gain of baroreflex parasympathetic control of HR | 5 | - |
| $K_{s,s}$ | Gain of baroreflex sympathetic control of aSKNA | 5 | - |
| $\tau_{vs}$ | Time constant of venous system dynamics | 0.5 | s |
| $\tau_{relax}$ | Coefficient describing input pressure on venous pressure | 1 | s |
| $\tau_r$ | Transition speed of ITP | 0.6 | s |
| $P_{vs,0}$ | Baseline venous pressure | 11.1 | mmHg |
| $k_r$ | Sympathetic effect on peripheral resistance | 0.1 | - |
| $P_b$ | Baseline arterial systolic blood pressure | 115 | mmHg |
| $C_{as}$ | arterial compliance | 1 | L/mmHg |
| $M_s$ | Sympathetic impact coefficient on HR | 0.3 | - |
| $M_t$ | Parasympathetic impact coefficient on HR | 0.5 | - |
| $\tau_{s,h}$ | Adaptation constant of sympathetic HR tone | 10 [29] | s |
| $\tau_{s,s}$ | Adaptation constant of sympathetic tone for aSKNA | 1 | s |
| $k_1$ | Coefficient related to baroreflex sensitivity | 1 | Hz/mmHg·s |
| $k_{SKNA}$ | Sympathetic impact coefficient on aSKNA | 10 | - |
| $\tau_p$ | Adaptation constant of parasympathetic HR tone | 1.8 [29] | s |

The model fitting performance were then evaluated for HR and aSKNA respectively based on three criteria: root mean square error (RMSE), mean absolute error (MAE), and Pearson correlation coefficient.

## III. Results

### A. Impact of different factors on mean iSKNA and HRV

Baseline characteristics of participants were described with mean ± standard deviation. Our population consisted of relatively young participants (Table II). A student’s t-test showed there is no significant difference on BMI between male and female subjects in this study (p=0.15).

**TABLE II.** Baseline characteristics of 41 subjects.

| Characteristics | Mean ± SD / n (%) |
| --- | --- |
| Sex [males] | 20 (49%) |
| Age [years] | 26.0 ± 10.5 |
| Height [m] | 178 ± 0.11 |
| Weight [kg] | 77.3 ± 15.1 |
| BMI [kg/m<sup>2</sup>] | 24.3 ± 3.8 |

Table III summarizes the LMM analysis results. A strong main effect of State was observed for mean iSKNA (F(1,234)=220.12, p<0.001), whereas the corresponding effect for RMSSD was weaker, although still statistically significant (F(1,234)=4.07, p=0.04). In addition, significant State × Sex and State × BMI interactions were present for iSKNA but not for RMSSD. As shown in the interaction plots (Fig. 3 (a)), the increase in mean iSKNA during VM was larger in male subjects. Fig. 3 (b) compares the estimate of subjects with different BMI (Low BMI = Mean(BMI) - SD(BMI), High BMI = Mean(BMI) + SD(BMI)). It can be observed the mean iSKNA estimate during VM was lower for subjects with higher BMI. No age-related effect was observed (Supplementary Material I), potentially due to only young adults were included. Additional results from all LMMs are provided in Supplementary Material I.

**TABLE III.** Linear mixed-effects model results for mean ISKNA and HRV (RMSSD)

| Effect | Mean iSKNA |  | HRV (RMSSD) |  |
| --- | --- | --- | --- | --- |
|  | F-statistic | p-value | F-statistic | p-value |
| State | F (1, 234) = 220.12 | <0.001 | F (1, 234) = 4.07 | 0.04 |
| BMI (centered) | F (1, 37.34) = 5.40 | 0.03 | F (1, 33.69) = 0.21 | 0.65 |
| Sex | F (1, 37.19) = 4.76 | 0.04 | F (1, 33.58) = 0.18 | 0.68 |
| State × Sex | F (1, 234) = 13.37 | <0.001 | F (1, 234) = 1.05 | 0.31 |
| State × BMI | F (1, 234) = 22.76 | <0.001 | F (1, 234) = 0.39 | 0.53 |

**Fig. 3.**
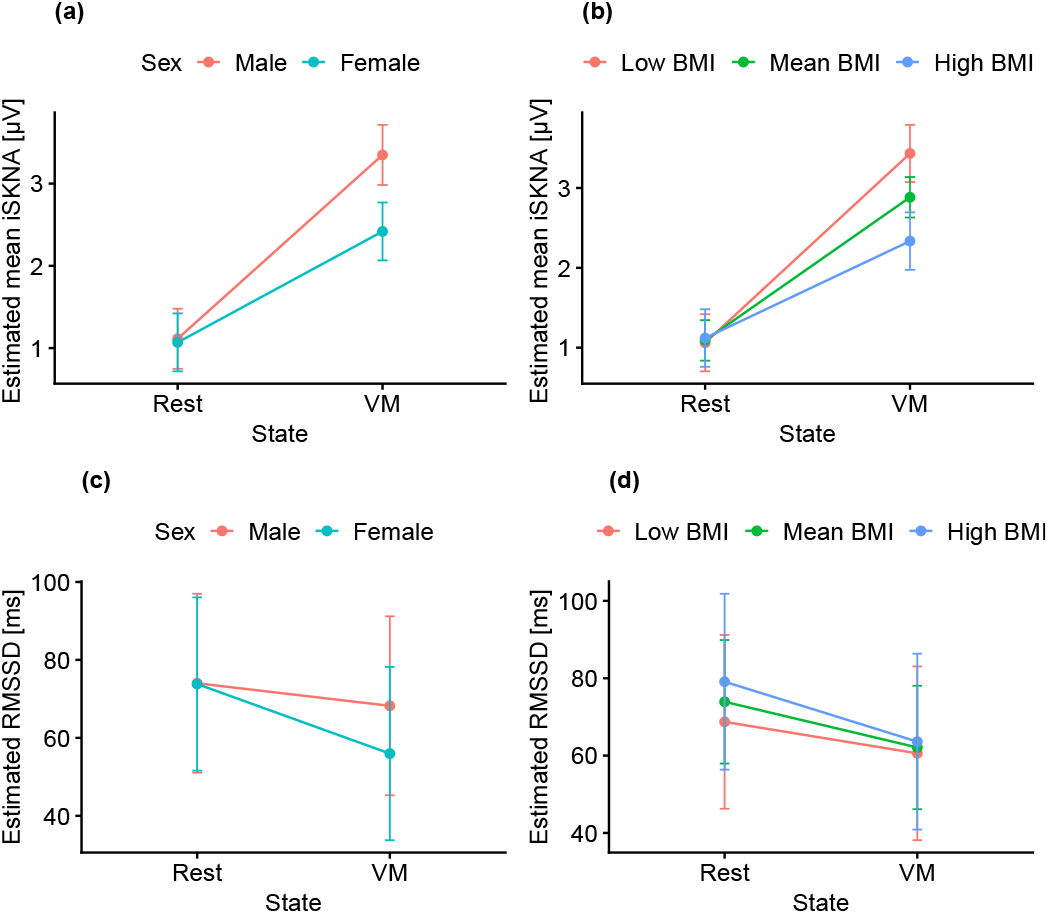
Interaction plots of subgroup-dependent condition effects. Statistical significance of main and interaction effects is reported in Table III. (a) and (b) show estimated mean iSKNA across rest and VM, stratified by sex and BMI levels (Low BMI = mean-SD; high BMI = mean+SD), respectively. (c) and (d) show the corresponding RMSSD estimates. Error bars represent 95% confidence intervals.

### B. Time–frequency analysis of respiratory coupling for iSKNA and RR intervals

Fig. 4 (a)-(b) illustrate the time-varying LF and HF band power for RR intervals and iSKNA. Given a window length of 25-s was used, the VM relevant segments extended from 47.5s–87.5s and 122.5s-162.5s. Both RR and iSKNA exhibited clear band power increases during the VM periods (indicated by the vertical lines). However, RR band power increased in a smooth and broad manner, consistent with the slowly varying nature of HRV. In contrast, iSKNA band power showed a more localized and sharper increase around the VM related period. In addition, the IQR area of HF power across both rest and VM periods was larger than LF for RR series. In contrast, HF power of iSKNA showed a larger IQR only at rest. Fig. 4 (c) showed the coherence between iSKNA and EDR. It can be observed that the mean HF coherence (red line) showed decreasing pattern during VM and increasing while transition to rest period, consistent with the breath holding movement. (d) shows coherence between RR series and EDR. HF coherence also showed a decrease pattern while transiting from rest to VM.

**Fig. 4.**
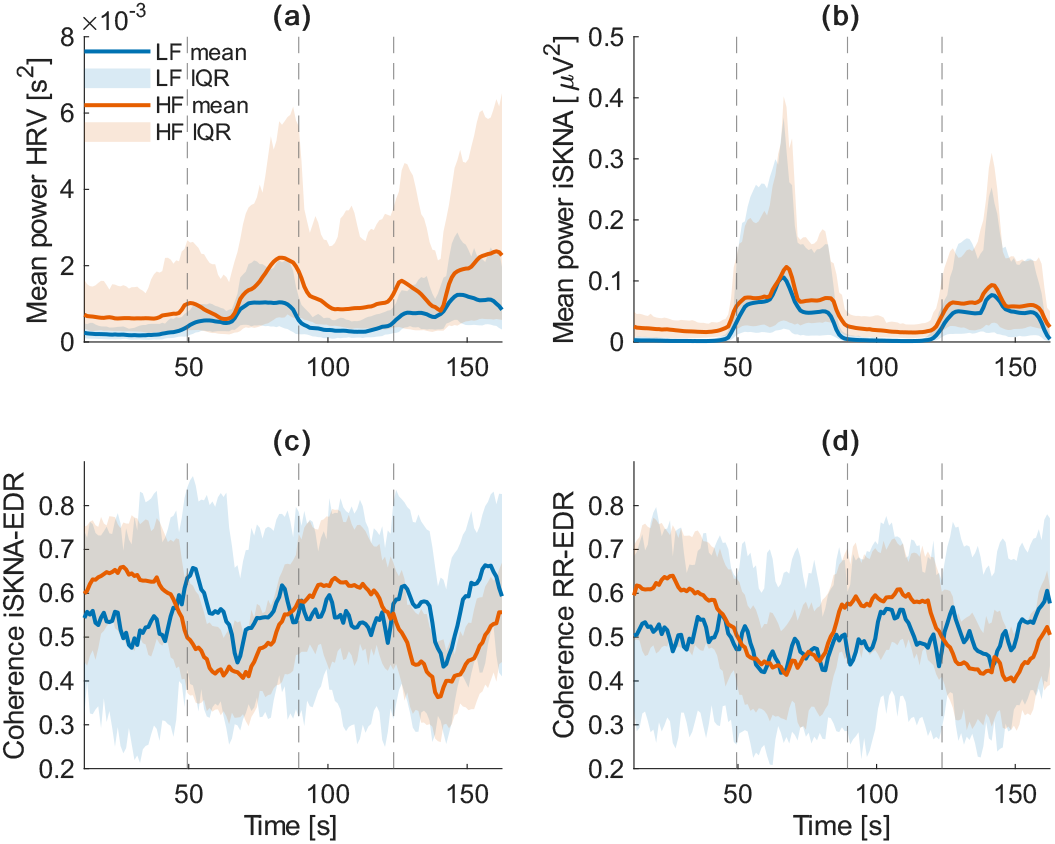
(a) Time-varying band power of HRV; (b) Time-varying band power of iSKNA; (c) Band spectral coherence between iSKNA and EDR; (d) Band spectral coherence between iSKNA and RR series. Blue line = low frequency; Red line = high frequency; Shaded areas = interquantile range. Dash lines labelled VM related periods (47.5s–87.5s, 122.5s–162.5s). Solid lines are 50% trimmed mean values and shaded area are 1^st^ to 3^rd^ quartile.

### C. Reproduction of HR and aSKNA by mathematical modeling

Table IV summarizes the results of the median and IQR of the estimated parameters. Most parameters exhibited relatively broad IQRs, reflecting large inter-subject and inter-session differences in VM responses, as well as the simplified nature of the model.

**TABLE IV.** Summary of the median and IQR of the identified parameters and their definitions, values and units.

| Parameters | Median [IQR] | Definition | Unit |
| --- | --- | --- | --- |
| $HR_b$ | 69.04 [17.61] | Baseline heart rate | beats/min |
| $aSKNA_b$ | 0.90 [0.27] | baseline aSKNA value | $\mu V$ |
| $R_0$ | 9.08 [15.87] | Baseline peripheral resistance | mmHg.s/L |
| $k_{vs}$ | 0.12 [0.08] | Impact of intra-thoracic pressure on venous pressure | - |
| $s_p$ | 0.48 [0.30] | Half-saturation value for parasympathetic HR control | - |
| $M$ | 147.53 [49.04] | Maximum baroreceptor firing rate | Hz |
| $k_{str}$ | 0.01 [0.003] | Coefficient describing heart contractility | mmHg/beat |
| $\tau_{resp,h}$ | 0.72 [2.15] | Adaptation constant of respiration dynamics for HR | s |
| $k_{resp,h}$ | 0.01 [0.06] | Respiratory impact coefficient on HR | - |
| $D_{resp,h}$ | 0.57 [0.98] | Delay for respiration mediated HR related parasympathetic tone | s |
| $\tau_{resp,s}$ | 0.25 [1.19] | Adaptation constant of respiration dynamics for aSKNA | s |
| $k_{resp,s}$ | 0.05 [0.15] | Respiratory impact coefficient on aSKNA | - |
| $D_{resp,s}$ | 0.81 [0.95] | Delay for respiration mediated aSKNA related sympathetic tone | s |

Fig. 5 (a) – (b) showed a representative example of model fitting. During VM, the aSKNA and HR elevation is largely reproduced by the model. During the resting periods, the respiratory modulation of both HR and aSKNA is also captured. The baroreflex-mediated sympathetic tone increases substantially during VM and gradually drops after the endpoint.

**Fig. 5.**
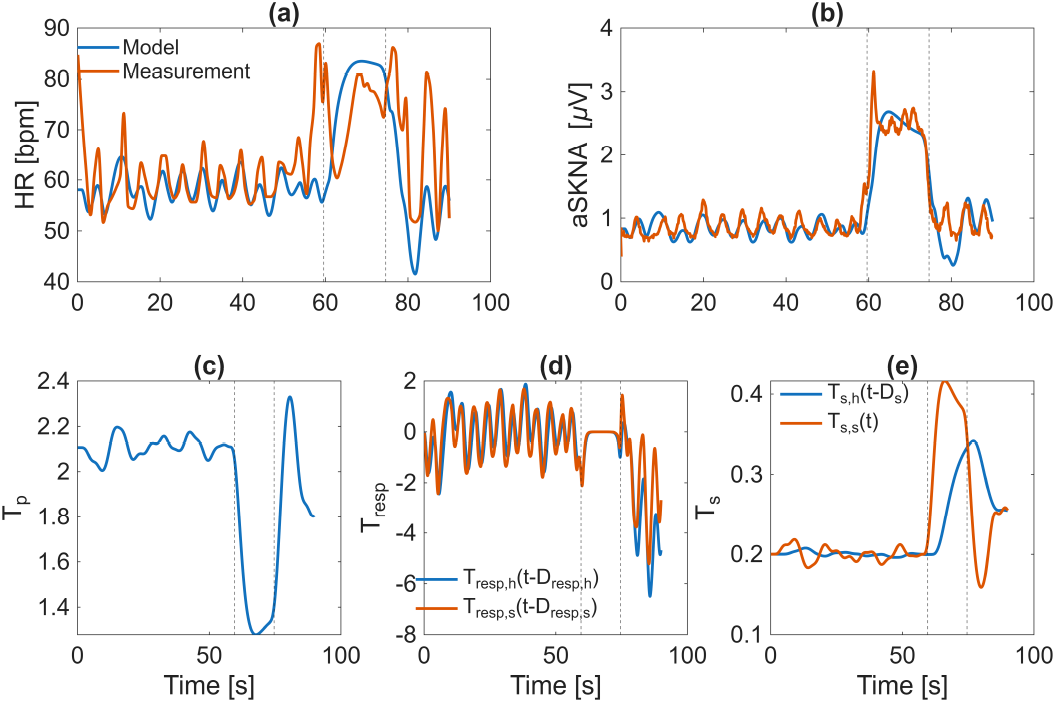
Example of model fitting results. (a) Red line: HR measurement; blue line: HR model estimation; (b) Red line: aSKNA measurement; blue line: aSKNA model estimation; (c) Baroreflex-mediated parasympathetic tone; (d) Red line: respiratory modulated aSKNA related sympathetic tone; Blue line: respiratory modulated HR related autonomic tone; (e) Red line: Baroreflex-mediated aSKNA related sympathetic tone; Blue line: Baroreflex-mediated HR related sympathetic tone. Dash lines labelled onset and offset of VM.

Overall, the model reconstructed aSKNA better than HR: the median RMSE for aSKNA was 0.42*µ*V with [Q_1_, Q_3_] = [0.27, 0.70], compared with 8.79bpm for HR with [Q_1_, Q_3_] = [6.15, 11.22]; the median MAE for aSKNA was 0.26*µ*V with [Q_1_, Q_3_] = [0.18, 0.41], compared with 6.01bpm for HR with [Q_1_, Q_3_] = [4.30, 7.61]; the median PCC for aSKNA was 0.80 with [Q_1_, Q_3_] = [0.60, 0.91], compared with 0.37 for HR with [Q_1_, Q_3_] = [0.16, 0.55], indicating better agreement.

## IV. Discussion

This study provides a complementary two-layer framework to assess the physiological interpretability of SKNA changes during autonomic challenges. We showed that iSKNA fluctuations at rest are predominantly driven by respiratory modulation, whereas during the VM they are mainly governed by baroreflex-mediated sympathetic activation. These observations were consistently supported by both time–frequency analysis and simplified physiological modeling. In addition, mean iSKNA showed a more pronounced response to VM than RMSSD in this cohort. We also observed that the iSKNA response varied with sex and BMI.

### A. Baroreflex impact on SKNA

The sympathetic activation during VM led to concomitant increases in HR and aSKNA, which were consistently reproduced by the model and reflected by elevated iSKNA as well as increased LF power in HRV (Fig. 4). This observation is in line with previous findings showing that SKNA is more sensitive than HRV for detecting sympathetic activation during autonomic challenges [5]. The baroreflex-related changes in HR exhibited different temporal characteristics than SKNA. While iSKNA responded rapidly and showed pronounced phase-dependent variations, RR-interval power increased more gradually (Fig. 1), reflecting the delayed sympathetic control of HR dynamics.

The model reproduced aSKNA dynamics more accurately than HR dynamics. Given the simplicity of the model, this difference suggests that aSKNA may offer a representation of sympathetic modulation that is more directly interpretable than HRV during VM. Consequently, SKNA may serve as a more interpretable indicator of sympathetic drive during VM in healthy subjects, particularly when rapid baroreflex responses are of interest.

### B. Respiratory modulation of SKNA

We presented that respiration, HR, and iSKNA exhibited a clear synchronous oscillatory pattern during rest, reflected by high HF coherence between EDR–iSKNA, and EDR–RR (Fig. 4). These findings suggest that iSKNA contains a stable respiratory-related modulation component, consistent with previous observations [7]. The relatively large HF IQR observed across subjects at rest further indicates considerable interindividual variability in respiratory-driven modulation and potentially reflecting differences in cardiorespiratory coupling level.

We further validated this modulation effect using a simple model. It turns out the respiratory modulation was well reproduced (Fig. 5), validating the high synchronization level between EDR and iSKNA. Together, these findings support the presence of a clear respiratory modulation of both RR and iSKNA under resting state in healthy subjects.

### C. Impact of Sex and BMI

In our healthy cohort, female participants showed a smaller increase in mean iSKNA during the VM than male participants. These findings partly matches previous research where women had lower baroreflex sensitivity [33], yet they mostly focused on middle aged subjects instead of younger cohorts in this study. Subjects with a higher BMI also showed a smaller mean iSKNA response during the VM, whereas no significant BMI-related difference was observed for RMSSD. This pattern suggests that mean iSKNA may be more sensitive than RMSSD in detecting between-subject variability in sympathetic responses during VM. It is known that Valsalva ratio has a negative correlation with BMI [34]. Since RMSSD primarily reflects vagal modulation, the discrepant findings between iSKNA and RMSSD may indicate that BMI had a greater influence on sympathetic rather than vagal aspects of the autonomic response.

Overall, these findings suggest that sex and BMI may influence the magnitude of SKNA responses and should be taken into account when interpreting SKNA amplitude as a quantitative biomarker of sympathetic tone. However, given the limited sample size and the relatively homogeneous healthy cohort, these findings should be considered preliminary. Although age was not significant in the present analysis, aging is known to substantially affect autonomic regulation and may contribute to additional variability in SKNA characteristics [35]. Future studies in larger and more heterogeneous cohorts are needed to further clarify these effects.

### D. SKNA as biomarker in clinical translation

Our findings about the influence of demographics, respiratory and baroreflex on SKNA indicate that its dynamics are strongly shaped by state-dependent modulations, underscoring the need for caution when interpreting SKNA amplitude as a biomarker of sympathetic tone across different conditions. SKNA has been increasingly used as a non-invasive surrogate of sympathetic activation, with multiple studies suggesting its potential for a wide range of clinical applications [2], [36]. Our results further support its higher responsiveness to acute autonomic challenges compared with conventional HRV indices.

### E. Limitations

This study has several limitations: First, the EDR is derived from a QRS area method, which is thought to capture respiration-related variations through electrical–mechanical coupling. However, its ability to reliably capture breath-holding phases during the VM has not been specifically validated. Therefore, conclusions drawn from EDR-based analysis during the VM phase should be interpreted with caution. Nevertheless, this method was selected because of its established performance in detecting respiratory disturbances such as sleep apnea [17]. Second, subject-performed VM may exhibit variable efforts, which likely contributed to the heterogeneity of HR and SKNA responses. The ITP was not strictly controlled in our protocol. Although a fixed 15-second window was used to locate the VM, the actual duration and intensity of the maneuver likely varied across trials and individuals, potentially contributing to inter-individual heterogeneity in HR and SKNA responses. Future studies incorporating direct measurements of airway pressure or respiratory effort could further standardize VM execution. Importantly, this methodological variability may coexist with the physiological mechanisms discussed above, as state-dependent respiratory and autonomic modulations can also influence SKNA expression during VM. Third, only 125 out of 158 VM sessions exhibited a significant SKNA burst, suggesting that in the remaining sessions the maneuver may not have been sufficiently performed or that bursts were not reliably detected. Second, the spectral analysis uses a 25- second window, which guarantees the spectral resolution of 0.04Hz. However, this low temporal resolution hampers the characterization of VM of 15 seconds. In addition, parameter identifiability in the proposed model was limited, likely due to the restricted amount of informative observation data during VM. As a result, several parameters that may vary across individuals were fixed, with inter-individual variability primarily reflected through parameters related to autonomic tone. Future studies incorporating multi-modal measurements, particularly continuous beat-to-beat BP, might enhance parameter identifiability. Such advancements would facilitate the independent validation of both the model structure and the resulting parameter estimates and may enable more precise, subject-specific physiological characterization. Finally, the present cohort consisted predominantly of younger, healthy individuals with a BMI<30 kg*/*m^2^. This restricted range of age and body composition may have reduced statistical power and limited the generalizability of our findings. Future studies involving larger and more heterogeneous populations are needed to determine whether aging significantly influences SKNA. In addition, relative fat mass could be incorporated in future analysis to replace BMI as it is a more accurate indicator of obesity [37].

## V. Conclusion

In this study, we investigated the physiological basis of SKNA change during VM by a dual-layer approach: observational signal analysis and mechanistic modeling. Time–frequency coherence revealed that iSKNA is governed by both respiratory modulation and baroreflex-mediated sympathetic recruitment. Besides, the mean iSKNA are more sensitive to charaterize the sympathetic activation during VM than conventional HRV indices. The model largely reproduced the characteristic VM-induced trajectory of aSKNA, providing supportive evidence that SKNA can reflect physiologically relevant sympathetic dynamics. Besides, we also found the mean iSKNA response during VM varies with sex and BMI levels.

## Data Availability

All data produced in the present study will be published at Zenodo in the future.

## Acknowledgment

The authors would like to acknowledge all students in technical medicine for data collection. This study is funded in part by the China Scholarship Council.

## Notes

### Competing Interest Statement

The authors have declared no competing interest.

### Funding Statement

This study did not receive any funding.

### Author Declarations

Initial data collection in healthy volunteers was approved by the Natural Science and Engineering Sciences Ethics Committee of the University of Twente (2022.153). Informed consent was signed by all volunteers. The reuse of the data for this study was approved by the Ethics Committee of the Computer and Information Sciences, University of Twente (nr. 230713).

## Reference

[1] T. Kusayama, J. Wong, X. Liu, W. He, A. Doytchinova, E. A. Robinson, D. E. Adams, L. S. Chen, S.-F. Lin, K. Davoren, R. G. Victor, C. Cai, M.-Y. Dai, Y. Tian, P. Zhang, D. Ernst, R. H. Rho, M. Chen, Y.-M. Cha, D. R. Walega, T. H. Everett, and P.-S. Chen, “Simultaneous noninvasive recording of electrocardiogram and skin sympathetic nerve activity (neuECG),” Nat Protoc, vol. 15, no. 5, pp. 1853–1877, May 2020. [Online]. Available: https://www.nature.com/articles/s41596-020-0316-6

[2] A. Doytchinova, J. L. Hassel, Y. Yuan, H. Lin, D. Yin, D. Adams, S. Straka, K. Wright, K. Smith, D. Wagner, C. Shen, V. Salanova, C. Meshberger, L. S. Chen, J. C. Kincaid, A. C. Coffey, G. Wu, Y. Li, R. J. Kovacs, T. H. Everett, R. Victor, Y.-M. Cha, S.-F. Lin, and P.-S. Chen, “Simultaneous noninvasive recording of skin sympathetic nerve activity and electrocardiogram,” Heart Rhythm, vol. 14, no. 1, pp. 25–33, Jan. 2017. [Online]. Available: https://www.sciencedirect.com/science/article/pii/S1547527116308025

[3] T. H. Everett, A. Doytchinova, Y.-M. Cha, and P.-S. Chen, “Recording sympathetic nerve activity from the skin,” Trends Cardiovasc. Med., vol. 27, no. 7, pp. 463–472, Oct. 2017. [Online]. Available: https://linkinghub.elsevier.com/retrieve/pii/S1050173817300579

[4] F. Baghestani, Y. Kong, W. D’Angelo, and K. H. Chon, “Analysis of sympathetic responses to cognitive stress and pain through skin sympathetic nerve activity and electrodermal activity,” Comput. Biol. Med., vol. 170, p. 108070, Mar. 2024. [Online]. Available: https://linkinghub.elsevier.com/retrieve/pii/S0010482524001549

[5] F. Baghestani, Y. Kong, A. V. Tolat, W. D’Angelo, and K. Chon, “Experimental Investigation of ECG-Derived Skin Nerve Activity in Sympathovagal Tone Assessment,” Rochester, NY, Sep. 2025. [Online]. Available: https://papers.ssrn.com/abstract=5518241

[6] W. He, Y. Tang, G. Meng, D. Wang, J. Wong, G. A. Mitscher, D. Adams, T. H. Everett, P.-S. Chen, and S. Manchanda, “Skin sympathetic nerve activity in patients with obstructive sleep apnea,” Heart Rhythm, vol. 17, no. 11, pp. 1936–1943, Nov. 2020. [Online]. Available: https://linkinghub.elsevier.com/retrieve/pii/S1547527120305968

[7] G. Meng, W. He, J. Wong, X. Li, G. A. Mitscher, S. Straka, D. Adams, T. H. Everett, S. Manchanda, X. Liu, P.-S. Chen, and Y. Tang, “Successful continuous positive airway pressure treatment reduces skin sympathetic nerve activity in patients with obstructive sleep apnea,” Heart Rhythm, vol. 19, no. 1, pp. 127–136, Jan. 2022. [Online]. Available: https://linkinghub.elsevier.com/retrieve/pii/S1547527121021664

[8] T. Kusayama, J. Wan, A. Doytchinova, J. Wong, R. A. Kabir, G. Mitscher, S. Straka, C. Shen, T. H. Everett, and P.-S. Chen, “Skin sympathetic nerve activity and the temporal clustering of cardiac arrhythmias,” JCI Insight, vol. 4, no. 4, p. e125853, Feb. 2019. [Online]. Available: https://insight.jci.org/articles/view/125853

[9] S. Chen, G. Meng, A. Doytchinova, J. Wong, S. Straka, J. Lacy, X. Li, P.-S. Chen, and T. H. Everett Iv, “Skin Sympathetic Nerve Activity and the Short-Term QT Interval Variability in Patients With Electrical Storm,” Front. Physiol., vol. 12, p. 742844, Dec. 2021. [Online]. Available: https://www.frontiersin.org/articles/10.3389/fphys.2021.742844/full

[10] J. L. Greaney and W. L. Kenney, “Measuring and quantifying skin sympathetic nervous system activity in humans,” J. Neurophysiol., vol. 118, no. 4, pp. 2181–2193, Oct. 2017. [Online]. Available: https://www.physiology.org/doi/10.1152/jn.00283.2017

[11] R. Lin, F. Halfwerk, D. Donker, G. D. Laverman, and Y. Wang, “A Mathematical Model for Skin Sympathetic Nerve Activity Simulation,” Aug. 2024, arXiv:2408.05934 [physics]. [Online]. Available: http://arxiv.org/abs/2408.05934

[12] J. Pan and W. J. Tompkins, “A real-time QRS detection algorithm,” IEEE Trans Biomed Eng, vol. 32, no. 3, pp. 230–236, Mar. 1985.

[13] T. Beauchaine, “Vagal tone, development, and Gray’s motivational theory: Toward an integratedmodel of autonomic nervous system functioning in psychopathology,” Dev. Psychopathol., vol. 13, no. 2, pp. 183–214, Jun. 2001.

[14] P. Grossman and K. Wientjes, “Respiratory Sinus Arrhythmia and Parasympathetic Cardiac Control: Some Basic Issues Concerning Quantification, Applications and Implications,” in Cardiorespiratory and Cardiosomatic Psychophysiology, P. Grossman, K. H. L. Janssen, and D. Vaitl, Eds. Boston, MA: Springer US, 1986, pp. 117–138. [Online]. Available: http://link.springer.com/10.1007/978-1-4757-0360-38

[15] C. Menuet, A. Ben-Tal, A. Linossier, A. M. Allen, B. H. Machado, D. J. A. Moraes, D. G. S. Farmer, D. J. Paterson, D. Mendelowitz, E. G. Lakatta, E. W. Taylor, G. L. Ackland, I. H. Zucker, J. P. Fisher, J. S. Schwaber, J. Shanks, J. F. R. Paton, J. Buron, K. M. Spyer, K. Shivkumar, M. Dutschmann, M. J. Joyner, N. Herring, P. Grossman, R. M. McAllen, R. Ramchandra, S. T. Yao, T. Ritz, and A. V. Gourine, “Redefining respiratory sinus arrhythmia as respiratory heart rate variability: an international Expert Recommendation for terminological clarity,” Nat Rev Cardiol, May 2025.

[16] G. B. Moody, R. G. Mark, A. Zoccola, and S. Mantero, “Derivation of respiratory signals from multi-lead ecgs,” Comput. Cardiol., vol. 12, no. 1985, pp. 113–116, 1985.

[17] N. Sadr and P. de Chazal, “A comparison of three ECG-derived respiration methods for sleep apnoea detection,” Biomed. Phys. Eng. Express, vol. 5, no. 2, p. 025027, Jan. 2019. [Online]. Available: 10.1088/2057-1976/aafc80

[18] J. Koenig and J. F. Thayer, “Sex differences in healthy human heart rate variability: A meta-analysis,” Neurosci. Biobehav. Rev., vol. 64, pp. 288–310, May 2016. [Online]. Available: https://www.sciencedirect.com/science/article/pii/S0149763415302578

[19] A. Voss, R. Schroeder, A. Heitmann, A. Peters, and S. Perz, “Short-Term Heart Rate Variability—Influence of Gender and Age in Healthy Subjects,” PLOS ONE, vol. 10, no. 3, p. e0118308, Mar. 2015. [Online]. Available: https://journals.plos.org/plosone/article?id=10.1371/journal.pone.0118308

[20] F. Shaffer and J. P. Ginsberg, “An Overview of Heart Rate Variability Metrics and Norms,” Front. Public. Health., vol. 5, p. 258, Sep. 2017. [Online]. Available: https://pmc.ncbi.nlm.nih.gov/articles/PMC5624990/

[21] D. Bates, M. Mächler, B. Bolker, and S. Walker, “Fitting linear mixed-effects models using lme4,” J. Stat. Softw., vol. 67, pp. 1–48, 2015.

[22] M. Orini, R. Bailon, L. T. Mainardi, P. Laguna, and P. Flandrin, “Characterization of Dynamic Interactions Between Cardiovascular Signals by Time-Frequency Coherence,” IEEE Trans. Biomed. Eng., vol. 59, no. 3, pp. 663–673, Mar. 2012. [Online]. Available: https://ieeexplore.ieee.org/document/6093939/

[23] M. Orini, P. Laguna, L. Mainardi, and R. Bailón, “Assessment of the dynamic interactions between heart rate and arterial pressure by the cross time–frequency analysis,” Physiol. Meas., vol. 33, no. 3, pp. 315–331, 2012.

[24] M. J. Prerau, R. E. Brown, M. T. Bianchi, J. M. Ellenbogen, and P. L. Purdon, “Sleep Neurophysiological Dynamics Through the Lens of Multitaper Spectral Analysis,” Physiology (Bethesda), vol. 32, no. 1, pp. 60–92, Jan. 2017. [Online]. Available: https://pmc.ncbi.nlm.nih.gov/articles/PMC5343535/

[25] L. Pstras, K. Thomaseth, J. Waniewski, I. Balzani, and F. Bellavere, “The valsalva manoeuvre: physiology and clinical examples,” Acta Physiol, vol. 217, no. 2, pp. 103–119, 2016.

[26] M. Kana and J. Holcik, “Mathematical model-based markers of autonomic nervous activity during the Valsalva Maneuver and comparison to heart rate variability,” Biomed. Signal Process. Control., vol. 6, no. 3, pp. 251–260, Jul. 2011. [Online]. Available: https://linkinghub.elsevier.com/retrieve/pii/S1746809411000413

[27] M. S. Olufsen, H. T. Tran, J. T. Ottesen, L. A. Lipsitz, and V. Novak, “Modeling baroreflex regulation of heart rate during orthostatic stress,” Am. J. Physiol. Regul. Integr. Comp. Physiol., vol. 291, no. 5, pp. R1355–R1368, Nov. 2006. [Online]. Available: https://www.physiology.org/doi/10.1152/ajpregu.00205.2006

[28] J. Ottesen and M. Olufsen, “Functionality of the baroreceptor nerves in heart rate regulation,” Comput. Methods Programs Biomed., vol. 101, no. 2, pp. 208–219, Feb. 2011. [Online]. Available: https://linkinghub.elsevier.com/retrieve/pii/S0169260710002749

[29] B. Randall, A. Billeschou, L. S. Brinth, J. Mehlsen, and M. S. Olufsen, “A model-based analysis of autonomic nervous function in response to the Valsalva maneuver,” J. Appl. Physiol., vol. 127, no. 5, pp. 1386–1402, Nov. 2019. [Online]. Available: https://www.physiology.org/doi/10.1152/japplphysiol.00015.2019

[30] K. Lu, J. W. Clark, F. H. Ghorbel, D. L. Ware, and A. Bidani, “A human cardiopulmonary system model applied to the analysis of the Valsalva maneuver,” Am. J. Physiol. Heart Circ. Physiol., vol. 281, no. 6, pp. H2661–H2679, Dec. 2001. [Online]. Available: https://www.physiology.org/doi/10.1152/ajpheart.2001.281.6.H2661

[31] F. Yasuma and J.-i. Hayano, “Respiratory sinus arrhythmia: why does the heartbeat synchronize with respiratory rhythm?” Chest, vol. 125, no. 2, pp. 683–690, 2004.

[32] J. J. Moré and D. C. Sorensen, “Computing a Trust Region Step,” SIAM J. Sci. and Stat. Comput., vol. 4, no. 3, pp. 553–572, Sep. 1983, tLDR: An algorithm for the problem of minimizing a quadratic function subject to an ellipsoidal constraint is proposed and it is shown that this algorithm is guaranteed to produce a nearly optimal solution in a finite number of iterations. [Online]. Available: http://epubs.siam.org/doi/10.1137/0904038

[33] H. V. Huikuri, S. M. Pikkujämsä, K. J. Airaksinen, M. J. Ikäheimo, A. O. Rantala, H. Kauma, M. Lilja, and Y. A. Kesäniemi, “Sex-related differences in autonomic modulation of heart rate in middle-aged subjects,” Circulation, vol. 94, no. 2, pp. 122–125, 1996.

[34] D. A. Gelber, M. Pfeifer, B. Dawson, and M. Schumer, “Cardiovascular autonomic nervous system tests: determination of normative values and effect of confounding variables,” J. Auton. Nerv. Syst., vol. 62, no. 1-2, pp. 40–44, 1997.

[35] K. Collins, A. Exton-Smith, M. James, and D. Oliver, “Functional changes in autonomic nervous responses with ageing,” Age Ageing, vol. 9, no. 1, pp. 17–24, 1980.

[36] J. Li and L. Zheng, “The Mechanism of Cardiac Sympathetic Activity Assessment Methods: Current Knowledge,” Front. Cardiovasc. Med., vol. 9, p. 931219, Jun. 2022. [Online]. Available: https://www.frontiersin.org/articles/10.3389/fcvm.2022.931219/full

[37] O. O. Woolcott and R. N. Bergman, “Relative fat mass (RFM) as a new estimator of whole-body fat percentage - a cross-sectional study in american adult individuals,” Sci. Rep., vol. 8, no. 1, p. 10980, 2018.

