## Supplementary Material for "Mechanistic Insights into Skin Sympathetic Nerve Activity Dynamics in Healthy Subjects Through a Two-Layer Signal-Analytical and Closed-Loop Physiological Modeling Framework"

### Supplementary Material I: LMM analysis for mean iSKNA and RMSSD

Before applying the linear mixed-effects model (LMM) used in the main text, several nested models were firstly examined to study the separately effects of State (Valsalva Maneuver vs. rest), sex, age, BMI, and interactions. These models are: $M_0$:

$$\text{F} \sim \text{State} + (1|\text{subject}) + (1|\text{subject} : \text{Measurement}) \quad (1)$$

$M_1$:

$$\text{F} \sim \text{Age} + (1|\text{subject}) + (1|\text{subject} : \text{Measurement}) \quad (2)$$

$M_2$:

$$\text{F} \sim \text{BMI} + (1|\text{subject}) + (1|\text{subject} : \text{Measurement}) \quad (3)$$

$M_3$:

$$\text{F} \sim \text{Sex} + (1|\text{subject}) + (1|\text{subject} : \text{Measurement}) \quad (4)$$

$M_4$:

$$\text{F} \sim \text{State}+\text{Sex}+(1|\text{subject})+(1|\text{subject} : \text{Measurement}) \quad (5)$$

$M_5$:

$$\text{F} \sim \text{State} + \text{BMI} + (1|\text{subject}) + (1|\text{subject} : \text{Measurement}) \quad (6)$$

$M_6$:

$$\begin{aligned}\text{F} \sim \text{State} + \text{Sex} + \text{BMI} + (1|\text{subject})\\ + (1|\text{subject} : \text{Measurement})\end{aligned} \quad (7)$$

$M_7$:

$$\begin{aligned}\text{F} \sim \text{State} + \text{Sex} + \text{BMI} + \text{State} \times \text{BMI}\\ + (1|\text{subject}) + (1|\text{subject} : \text{Measurement})\end{aligned} \quad (8)$$

$M_8$:

$$\begin{aligned}\text{F} \sim \text{State} + \text{Sex} + \text{BMI} + \text{State} \times \text{Sex}\\ + (1|\text{subject}) + (1|\text{subject} : \text{Measurement})\end{aligned} \quad (9)$$

Specifically, $M_0$ examined the State effect alone; $M_1$–$M_3$ examined the separate effects of age, BMI, and sex; $M_4$–$M_6$ added covariates to the State model; and $M_7$–$M_8$ further examined State interaction terms. The models were tested with type III F-test with Satterthwaite's method. Table I summarizes the results.

Overall, the nested models showed a robust VM-induced effect on mean iSKNA and a weaker effect on RMSSD. Sex, BMI, and their interactions with State were associated with mean iSKNA, whereas no significant interaction effects were observed for RMSSD. These findings support the inclusion of sex- and BMI-related terms in the main LMM analysis.

### Supplementary Material II: Model parameter estimation

#### A. Calculation of $k_{str}$

$k_{str}$ is the coefficient that relates to cardiac contractility at rest where there is no SBP change, according to Eq. 3 in the main text:

$$\frac{dP_{as}(t)}{dt} = \frac{1}{C_{as}}[Q_{in}(t) - Q_{out}(t)] = 0 \quad (10)$$

Runwei Lin is with the Department of Biomedical Signals and Systems, University of Twente, Enschede, the Netherlands.
Frank R. Halfwerk is with the Cardiac Surgery Innovations Lab, Engineering Organ Support Technologies Group, Department of Biomechanical Engineering, University of Twente, Enschede, the Netherlands, and with the Department of Cardio-thoracic surgery, Thorax Centrum Twente, Medisch Spectrum Twente, Enschede, the Netherlands, and with the Cardiovascular Health Technology Centre, TechMed Centre, University of Twente, the Netherlands.
Dirk W. Donker is with the Cardiovascular and Respiratory Physiology Group, University of Twente, Enschede, the Netherlands, and with Intensive Care Center, University Medical Center Utrecht, Utrecht, the Netherlands.
Jacomine Tertoolen is with Engineering Organ Support Technologies Group, Department of Biomechanical Engineering, University of Twente, Enschede, the Netherlands, and with the Department of Cardio-thoracic surgery, Thorax Centrum Twente, Medisch Spectrum Twente, Enschede, the Netherlands.
V.R. van der Pas is with the Department of Cardio-thoracic surgery, Thorax Centrum Twente, Medisch Spectrum Twente, Enschede, the Netherlands.
Gozewijn Dirk Laverman is with the Department of Biomedical Signals and Systems, University of Twente, and with the Department of Internal Medicine, Ziekenhuisgroep Twente, Almelo, the Netherlands.
Ying Wang is with the Department of Biomedical Signals and Systems, University of Twente, and with Cardiovascular Health Technology Centre, TechMed Centre, University of Twente, the Netherlands.

TABLE I
TYPE III F-TEST RESULTS FROM NESTED LINEAR MIXED-EFFECTS MODELS FOR MEAN ISKNA AND RMSSD.

| Model | Effect | Mean iSKNA | | HRV (RMSSD) | |
|---|---|---|---|---|---|
| | | F-statistic | p-value | F-statistic | p-value |
| $M_0$ | State | F(1, 236) = 186.89 | < 0.001 | F(1, 236) = 4.24 | 0.04 |
| $M_1$ | Age | F (1, 36.77) = 1.13 | 0.29 | F (1, 32.45) = 2.93 | 0.10 |
| $M_2$ | BMI | F (1, 38.84) = 5.68 | 0.02 | F (1, 34.22) = 0.19 | 0.67 |
| $M_3$ | Sex | F (1, 37.20) = 5.06 | 0.03 | F (1, 34.16) = 0.15 | 0.70 |
| $M_4$ | State | F (1, 236) = 186.89 | < 0.001 | F (1, 236) = 4.24 | 0.04 |
| | Sex | F (1, 34.20) = 5.06 | 0.03 | F (1, 34.16) = 0.15 | 0.70 |
| $M_5$ | State | F (1, 236) = 186.89 | < 0.001 | F (1, 236) = 4.24 | 0.04 |
| | BMI | F (1, 38.84) = 5.68 | 0.02 | F (1, 34.21) = 0.18 | 0.68 |
| $M_6$ | State | F (1, 236) = 186.89 | < 0.001 | F (1, 236) = 4.24 | 0.04 |
| | BMI | F (1, 34.34) = 5.40 | 0.03 | F (1, 33.69) = 0.21 | 0.65 |
| | Sex | F (1, 37.19) = 4.76 | 0.04 | F (1, 33.59) = 0.18 | 0.68 |
| $M_7$ | State | F (1, 235) = 205.50 | < 0.001 | F (1, 235) = 4.23 | 0.04 |
| | BMI | F (1, 37.34) = 5.40 | 0.03 | F (1, 33.69) = 0.21 | 0.65 |
| | Sex | F (1, 37.19) = 4.76 | 0.04 | F (1, 33.59) = 0.18 | 0.68 |
| | State × BMI | F (1, 235) = 24.50 | < 0.001 | F (1, 235) = 0.50 | 0.48 |
| $M_8$ | State | F (1, 235) = 201.86 | < 0.001 | F (1, 235) = 4.07 | 0.04 |
| | BMI | F (1, 37.34) = 5.40 | 0.03 | F (1, 33.69) = 0.21 | 0.65 |
| | Sex | F (1, 37.19) = 4.76 | 0.04 | F (1, 33.59) = 0.18 | 0.68 |
| | State × Sex | F (1, 235) = 15.01 | < 0.001 | F (1, 235) = 1.17 | 0.28 |

we have $Q_{in}(t) = Q_{out}(t)$, and $P_{as}(t) = P_b$, $\text{HR}(t) = \text{HR}_b$, $R(t) = R_0$ and $P_{vs}(t) = P_{vs,0}$. From Eq. 5-8 in the main text we can derive

$$\frac{\text{HR}_b \cdot k_{str} \cdot P_{vs,0}}{P_b} = \frac{(P_b - P_{vs,0})}{R_0} \tag{11}$$

As a result, $k_{str}$ is defined as

$$k_{str} = \frac{P_b \cdot (P_b - P_{vs,0})}{\text{HR}_b \cdot R_0 \cdot P_{vs,0}} \tag{12}$$

#### B. Calculation of $s_p$ and $s_{s,h}$, and $s_{s,s}$

The $s_p$ and $s_{s,h}$, and $s_{s,s}$ are coefficients related to autonomic tone calculation. We followed the formulation described in [1] to calculate $s_p$, $s_{s,h}$, and $s_{s,s}$. A sympathetic-parasympathetic ratio of 0.2: 0.8 was assumed in resting period where $\text{HR}(t) = \text{HR}_b$. Consequently, the half-saturation values were obtained as:

$$s_p = \bar{n} + \frac{1}{q_p} \ln\left(\frac{K_p}{\bar{T}_p} - 1\right) \tag{13}$$

$$s_{s,h} = \bar{n} - \frac{1}{q_s} \ln\left(\frac{K_{s,h}}{\bar{T}_{s,h}} - 1\right), \tag{14}$$

$$s_{s,s} = \bar{n} - \frac{1}{q_s} \ln\left(\frac{K_{s,s}}{\bar{T}_{s,s}} - 1\right), \tag{15}$$

where $\bar{n} = N_0/M$, $\bar{T}_p = 0.8$, $\bar{T}_{s,h} = \bar{T}_{s,s} = 0.2$.

### SUPPLEMENTARY MATERIAL III: SENSITIVITY ANALYSIS AND PARAMETER IDENTIFICATION

A local sensitivity analysis was performed on the 24 parameters to determine the identifiable parameters in the model during VM. Fig. 1 showed the process for selecting identifiable parameters. To calculate the local sensitivity, the residual vector $\mathbf{R}(\theta_i)$ is firstly defined as stacking the normalized HR and aSKNA residual:

$$\mathbf{R}(\theta_i) = [r_{\text{HR}}(t_j), r_{\text{aSKNA}}(t_j)]^T, j = 1, 2, ..., N \tag{16}$$

where

$$r_{\text{HR}}(t_j) = \frac{\text{HR}(t_j) - \overline{\text{HR}_M}(t_j)}{\text{HR}_b{}^2} \tag{17}$$

$$r_{\text{aSKNA}} = \frac{\text{aSKNA}(t_j) - \overline{\text{aSKNA}_M}(t_j)}{\text{aSKNA}_b} \tag{18}$$

$\text{aSKNA}_b(t_j)$ and $\text{HR}_b(t_j)$ denote the baseline HR and baseline aSKNA. $\overline{\text{aSKNA}_M}(t_j)$ and $\overline{\text{HR}_M}(t_j)$ denote HR and aSKNA measurements.

To account for differences in parameter magnitudes, a natural logarithmic transformation was applied to each parameter. The local sensitivity of parameter $\theta_i$ was was then computed using a central finite-difference approximation:

$$\mathbf{J}_i = \frac{\mathbf{R}(ln\theta_i + h\mathbf{e}_i) - \mathbf{R}(ln\theta_i - h\mathbf{e}_i)}{2h}, h << 1 \tag{19}$$

The integration tolerance was set at $\phi = 10^{-8}$, and a sensitivity threshold of $10\sqrt{\phi} = 10^{-3}$ was adopted, consistent with previous studies [1], [2]. The local sensitivity for $\theta_i$ was defined as the $L_2$-norm of $i$th column of $S$.

$$s_i = ||\mathbf{J}_i||_2 \tag{20}$$

All local sensitivities were normalized. Following [1], we discarded parameters with normalized sensitivity below $10^{-3}$.

The SVD–QR method, a widely used approach for selecting identifiable parameter subsets, was applied to identify parameters suitable for estimation. Specifically, the Fisher information matrix (FIM) was decomposed to by singular value decomposition determine its numerical rank, defined as the number of singular values exceeding 0.001. This rank reflects the number of parameters that can be reliably identified from the data. Based on the estimated rank, QR decomposition with column pivoting was then applied to the sensitivity matrix to select the corresponding subset of parameters. Consequently, a subset of five parameters selected: $\{M, k_{vs}, s_p, R_0, k_{\text{SKNA}}\}$.

To further validate the parameter identifiability of the chosen subset, a structured correlation analysis was performed [1], [2]. This method evaluates pairwise correlations between parameters to identify combinations that cannot be reliably estimated simultaneously. Let $\mathbf{C}$ denotes the pseudoinverse matrix of FIM $F_M$. The correlation matrix was then defined as:

$$c_{ij} = \frac{\mathbf{C}_{ij}}{\sqrt{\mathbf{C}_{ii}\mathbf{C}_{jj}}} \tag{21}$$

The resulting correlation matrix is shown in Fig. 2 (b). Parameters were considered jointly identifiable if $|c_{ij}| < 0.9$. Since the structural correlation between $M$ and $k_{\text{SKNA}}$ exceed the threshold, and M presented a higher sensitivity, $k_{\text{SKNA}}$ was from the estimation process. The remaining four parameters were therefore retained for optimization, whereas all other parameters were fixed. Consequently, the selected parameter set was:

$$\theta_{sub} = \{M, k_{vs}, s_p, R_0\} \tag{22}$$

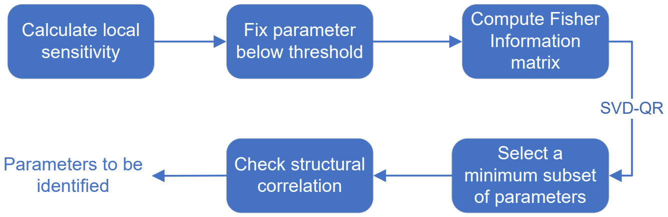


Fig. 1. Workflow for selecting identifiable parameters.

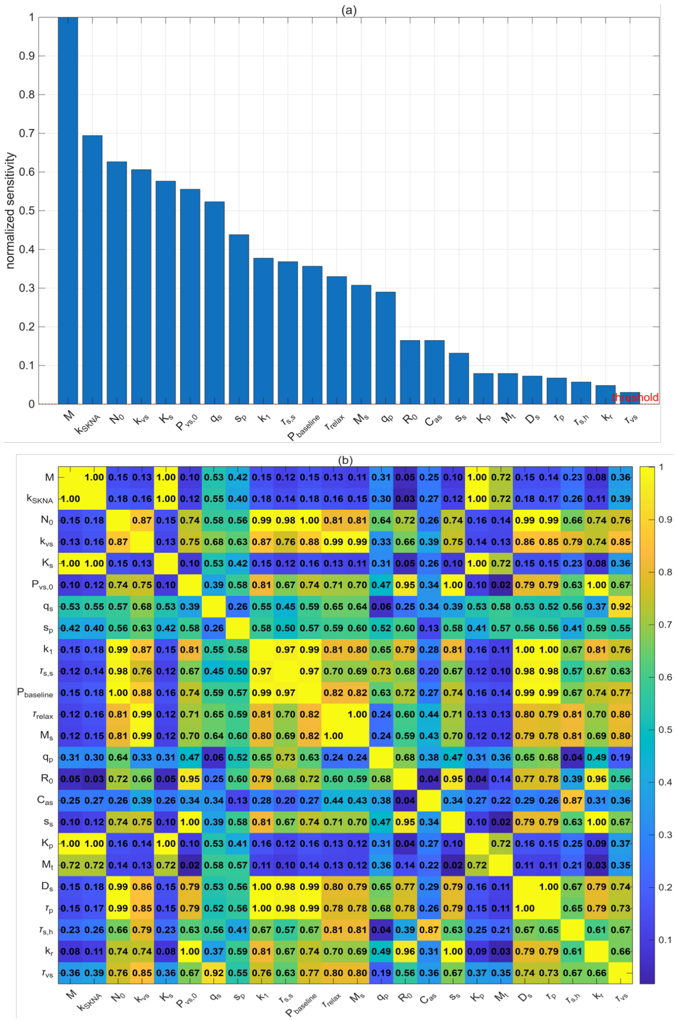


Fig. 2. Results of parameter identification analysis. (a) Normalized local sensitivity indices of the parameters. The red dashed line denotes the predefined sensitivity threshold. (b) Absolute values of the structural correlation matrix.

### REFERENCE


[1] E. B. Randall, A. Billeschou, L. S. Brinth, J. Mehlsen, and M. S. Olufsen, "A model-based analysis of autonomic nervous function in response to the Valsalva maneuver," *J. Appl. Physiol.*, vol. 127, no. 5, pp. 1386–1402, Nov. 2019. [Online]. Available: https://www.physiology.org/doi/10.1152/japplphysiol.00015.2019

[2] T. Wang, J. Wu, F. Qin, H. Jiang, X. Xiao, and Z. Huang, "Computational modeling for the quantitative assessment of cardiac autonomic response to orthostatic stress," *Physiol. Meas.*, vol. 45, no. 7, p. 075009, Jul. 2024. [Online]. Available: https://doi.org/10.1088/1361-6579/ad63ee
